# Temperature modulation of microvascular, inflammatory and perceptual responses to mechanical loading of the skin in young and older adults and in spinal cord injury patients

**DOI:** 10.64898/2026.07.14.26358023

**Authors:** Charlotte Stevens, Ralph Gordon, Sara Bergstrand, Amanda Feldt, Denisse Marginean, Bijar Ghafouri, Peter Worsely, Davide Filingeri

**Affiliations:** Thermosenselab, Skin Sensing Research Group, School of Health Sciences, University of Southampton, United Kingdom; PressureLab, Skin Sensing Research Group, School of Health Sciences, University of Southampton, United Kingdom; Department of Health, Medicine, and Caring Sciences, Linköping University, Linkoping, Sweden; PAINOMICS Research Group, Department of Health, Medicine and Caring Sciences, Linköping University, Sweden; Tyndall Centre for Climate Change Research, University of Southampton, United Kingdom

**Keywords:** skin temperature, pressure ulcers, cold temperature, spinal cord injuries, aging, biophysics, tissue viability, perception

## Abstract

Cooling the skin may increase its tolerance to mechanical loading and decrease the risk of developing pressure ulcers. Yet, the mechanisms of action (e.g. cooling-modulation of cytotoxic, post-occlusive hyperaemia), and their individual variability, remain unclear. We investigated the effects of different cooling levels (24°C and 16°C) on microvascular, inflammatory and perceptual responses to mechanical loading of the sacrum in healthy young (N=23) and older adults (N=19), and in spinal cord injury patients (SCI; N=10). Healthy participants underwent 45-min loading (∼60 mmHg) and 20-min unloading of the sacrum, using an instrumented indenter probe set at either 38°C (control condition), 24°C or 16°C. SCI participants completed a more conservative protocol (i.e. 25min, ∼45mmHg loading, 38°C and 16°C conditions). Pre-insult skin structure was characterised with optical coherence tomography; skin blood flow (SkBF) at the loading site was continuously measured, alongside thermal acceptability; and post-insult inflammatory responses were determined via skin-sebum cytokines analyses. Compared to control, 24°C- and 16°C-cooling induced a similar ∼8-fold decrease in peak post-occlusive reactive hyperaemia in healthy participants, with similar temperature-related differences observed in SCI. Pro-inflammatory cytokines decreased post-insult; yet this occurred similarly across all temperatures and groups. The majority of participants (≥70%) rated both 24°C- and 16°C-cooling as thermally acceptable. We conclude that cooling is a potent modulator of the skin microvascular response to mechanical loading in younger, older, and vulnerable skin (SCI). These findings can inform design parameters for thermal technology aimed at preventing the loss of skin integrity (e.g. integrating 24°C-cooling in support surfaces and skin wearables).

## Introduction

Pressure injuries/ulcers are localized damage to the skin and/or underlying tissue, usually over a bony prominence, resulting from prolonged pressure or pressure in combination with shear (Kottner *et al*., 2025). Exposure to prolonged mechanical loading of the skin can arise from several situations, including periods in lying and sitting postures (e.g. consider hospitalization), as well as the prolonged attachment of medical devices, e.g. prosthetics or respiratory masks (Kottner *et al*., 2018, 2025). In the United Kingdom alone, the annual cost of treating chronic wounds, including PU, has been estimated to be approximately £8.3 billion per annum (Guest *et al*., 2015). An improved understanding of the physiological mechanisms that determine skin tissue tolerance to sustained mechanical loading may therefore inform the development of cost-effective, personalised strategies to prevent pressure ulcers (PU) and improve patient care and quality of life (Roussou *et al*., 2023).

The pathophysiology of PU is multifactorial, involving both intrinsic (age, nutrition, skin status, metabolic supply and demand) and extrinsic factors (skin moisture, friction and shear) (Coleman *et al*., 2014). Skin deformations resulting from loading and shearing forces leads to internal tissue deformations, causing changes to the physiology of skin and sub-dermal tissue, including ischemia in the blood vasculature, lymphatic impairment, and direct deformation damage (Kottner *et al*., 2025). When the load is removed, post-occlusive reactive hyperaemia occurs, causing ischaemia-reperfusion injury through the release of oxygen-derived free radicals with cytotoxic effects (Tzen *et al*., 2013). In addition, microclimate conditions within and around skin tissues strongly influence the tolerance of skin to mechanical loading and shear. As an example, elevated temperature and humidity at the skin interface reduce its mechanical stiffness and increases the friction coefficient during shearing (Kottner *et al*., 2018). These mechanisms can lead to a greater risk of tissue damage for the same mechanical stress.

In contrast, theoretical modelling studies have indicated that lowering human skin temperature from 34 to 25 °C could provide a level of protection equivalent to that produced by a 30% reduction in mechanical loading (Lachenbruch, 2005). Evidence from animal models underpins the idea that the risk of PU formation can be reduced by lowering skin temperature (Filingeri *et al*., 2021). Using a rat skin model, local cooling has been shown to protect against loading-induced ischaemia (Jan *et al*., 2012; Lee *et al*., 2014). Similarly, in a porcine model, improved tissue integrity was observed with decreases in temperature to 25 °C, during 5-h mechanical loading equivalent to 100mmHg of pressure (Kokate *et al*., 1995). Histological examination of the tissue exposed to 25 °C revealed no soft tissue damage, whereas elevated temperatures at ≥35 °C resulted in significant damage. Preliminary animal models have also indicated that local cooling, as well as the stimulation of cold sensitive TRPM8-expressing neurons in dorsal root ganglions, could modulate the skin’ inflammatory responses to acute mechanical stress (e.g. loading) (Lee *et al*., 2014) and chronic states (e.g. chronic dermatitis) (Wang *et al*., 2020), via downregulation of the expression of pro-inflammatory cytokines such Tumour Necrosis Factor TNF-α.

The evidence above indicates that skin cooling could be a promising preventative intervention for maintaining skin integrity under mechanical loading; yet, its mechanisms of action in human skin *in vivo* remain poorly understood. Specifically, it remains unclear whether there is an optimal level of skin cooling (i.e. more or less pronounced) that can offer protection, and to what extent this effect arises from a combination of a) preserved microvascular perfusion during mechanical loading; b) attenuated post-reactive hyperaemia following mechanical unloading; and c) downregulation of skin’s inflammatory responses to sustained mechanical loading and unloading.

In addition to its potential physiological effects, it is well known that localised cooling of the skin induces cold discomfort, which greatly limits acceptability and adherence to interventions designed to maintain skin health, particularly for vulnerable individuals at risk of PUs such as the elderly (Ledger *et al*., 2020). Yet, there is limited evidence on the relationship between the physiological and perceptual effects of varying levels of skin cooling (i.e. approaching the cold-pain threshold of ∼15 °C skin temperature) during mechanical loading (Filingeri *et al*., 2014).

It should also be recognised that the physiological and perceptual effects of cooling to loading-induced damage are likely to vary as a function of age and comorbidities. These states are associated with changes in skin biophysics and morphology (Holbrook & Odland, 1974), and in thermoregulatory and perceptual sensitivities. For example, ageing is likely to modulate the effects of cooling on tissue tolerance, as aged skin presents a reduced physiological and perceptual sensitivity to cold, due to decreases in both reflex cutaneous vasoconstriction, and density of thermoreceptors (Johnson *et al*., 2014a). Similarly, the presence of a spinal cord injury (SCI) is associated with autonomic [e.g. impaired control of skin blood flow (SkBF)] and sensory disfunctions (e.g. perceptual loss below injury level) (Zeilig *et al*., 2012; Forsyth *et al*., 2019). Thus, there may be variations between reductions in cold sensitivity and diminished efficacy of cooling associated with age and clinical status.

Identifying how and why the effects of localised cooling vary both physiologically and perceptually according to age and clinical status, is critical to develop “user-centred” preventative approaches that are both effective and comfortable for groups at greater risk of PU formation, such as the elderly and those with a SCI. Indeed, several support surfaces and therapeutic interfaces (e.g. hospital mattresses and wheelchair cushions) already integrate microclimate management systems, despite mechanistic, empirical evidence of their efficacy remains limited (Van Leen *et al*., 2017; Worsley & Bader, 2019; Denzinger *et al*., 2019).

The aim of this study was to investigate how varying levels of localised cooling (i.e. 24 and 16 °C) modulate the microvascular, inflammatory, and perceptual responses to mechanical loading and unloading of the skin in young and older adults, and in a sub-cohort of patients at high risk of PU, i.e. those with a SCI. We hypothesised that skin cooling would downregulate both microvascular (e.g. peak post-occlusive reactive hyperaemia) and the inflammatory responses to mechanical loading, while potentially upregulating discomfort, in a dose-dependent fashion. We also hypothesised that these responses would be attenuated in aged skin and in the presence of SCI.

## Methods

### Ethical approval

All participants provided written informed consent before taking part in this study and were provided with the opportunity to withdraw at any time. The study adhered to the guidelines set out by the Declaration of Helsinki except for registration in a database. All experimental procedures were approved by the Ethics Committee at the University of Southampton (ERGO 88984) and by the UK Heath Research Authority (IRAS 350078).

### Participants

To establish how localised cooling modulated the microvascular, inflammatory and perceptual responses to mechanical loading and unloading in aged and vulnerable skin, we recruited 3 cohorts of participants, namely i) younger healthy adults (i.e. 18 – 35 years old), ii) older healthy adults (i.e. 55 – 70 years), and iii) individuals with a SCI (injury level T1 – S1), according to the following criteria:

#### Inclusion criteria

1. 18-70 years old;
2. Male or female;
3. Skin tone dark, medium, or light (assessed via the Fitzpatrick scale);
4. Healthy groups only: Physically active (i.e. performing exercise1 to 3 times a week);
5. SCI group only: presenting thoracic injury/paraplegia (i.e. injury level within T1-T12), >12 months post-injury, and having a history of PU;

#### Exclusion criteria

1. Healthy groups only: suffering from cardiovascular, metabolic, and neurological disorders and/or co-morbidities, e.g. hypertension, diabetes, chronic lung disease; Raynaud’s disease; and from skin conditions (e.g. eczema)
2. Healthy groups only: under drug therapy affecting thermoregulation (e.g. muscarinic antagonists); and active smoker or vaper;
3. SCI group only: <12 months post-injury and presenting a PU at the sacrum;

Changes in peak post-occlusive reactive hyperaemia were selected as the primary experimental outcome, as this represents a robust and reasonably repeatable microvascular response under well-controlled conditions, and that is directly implicated in the pathophysiology of PU (Agarwal *et al*., 2009; Liao *et al*., 2013). Using published data on the mean changes in peak post-occlusive reactive hyperaemia between a nerve block and control conditions (Lorenzo & Minson, 2007), we performed a sample size calculation (i.e. effect size f=0.4, α = 0.05; β = 0.80; between temperature conditions comparison) to determine a minimum sample size of 18 participants per group. The sample size calculation was used to inform the recruitment of the healthy cohorts (and subsequent analysis – see “Data and statistical analyses” section); whereas a convenience sampling approach was used to recruit the SCI group, owing to well-known clinical challenges associated with recruitment and testing in this group.

Following recruitment and screening, a total of 52 participants took part in the study, divided into the healthy younger (N=23) and older (N=19) groups, and the SCI group (N=10). Participants characteristics are described in Table 1.

**Table 1:** Summary of participant characteristics in the three experimental groups. Data presented as Mean ± SD or ratios where applicable.

| Variable | Young Healthy (n = 23) | Older Healthy (n = 19) | Spinal Cord Injured (n = 10) |
| --- | --- | --- | --- |
| Age (yrs) | 24 ± 4 | 66 ± 4 | 57 ± 10 |
| Mass (kg) | 71.1 ± 9.1 | 70.5 ± 14.3 | - |
| Height (cm) | 175.6 ± 8.5 | 170.7 ± 9.7 | - |
| Skin tone ratio<br>(light:medium:dark) | 8:12:3 | 7:12:0 | 3:7:0 |
| Spinal Injury type ratio<br>(complete:incomplete) | - | - | 5:5 |
| Male:Female | 10:13 | 8:11 | 6:4 |

### Experimental design

This study adopted a randomised cross-over design. A detailed account of the study design and protocol has been published previously, and it is available open access (Gordon *et al*., 2024).

Briefly, the healthy participants groups (younger and older) were exposed to an experimental protocol that delivered sustained mechanical loading (i.e. ∼60mmHg of pressure for 45 min) and unloading (i.e. ∼17 mmHg of pressure for 20 min) of the skin over the sacrum, using a custom-built thermo-mechanical indenter probe (i.e. 25-cm^2^ square surface), to induce skin tissue ischaemia and post-occlusive reactive hyperaemia. The protocol comprised three different probe temperatures, i.e. 38 °C (control - no cooling), 24 °C (mild cooling) or 16 °C (intense cooling), which were used in each of 3 separate testing sessions on different days, separated by a minimum of 24h. The protocol lasted 75 minutes in total and it consisted of the following:

i) a 10-minute baseline period with minimal loading (i.e. tare pressure of the probe sitting on participants’ skin= ∼17mmHg) at neutral temperature (i.e. 34 °C)
ii) a 5-minute loading period with no temperature manipulation (i.e. 34 °C), with a target load of ∼60mmHg, which was individually determined pre-loading, using a pressure mapping device
iii) a 40-minute loading period with temperature manipulation (i.e. either 38, 24 or 16 °C), with a target loading of ∼60 mmHg
iv) a 20-minute unloading phase with minimal loading (i.e. tare pressure of ∼17mmHg and maintained temperature manipulation of either 38, 24 or 16 °C)

Continuous force and temperature measurements during the protocol were possible due to the integration of force and temperature sensors into the thermo-mechanical probe, which sampled data at 30 Hz during all testing (DasyLab Version 2016 14.0.0). Average force and temperature data over the 75-min protocol are presented in **Figure 1A and B** for the healthy cohorts (note when considering Figure 1: the force measurements was zeroed during the 10-minute baseline period with minimal loading to reflect tare weight).

**Figure 1:**
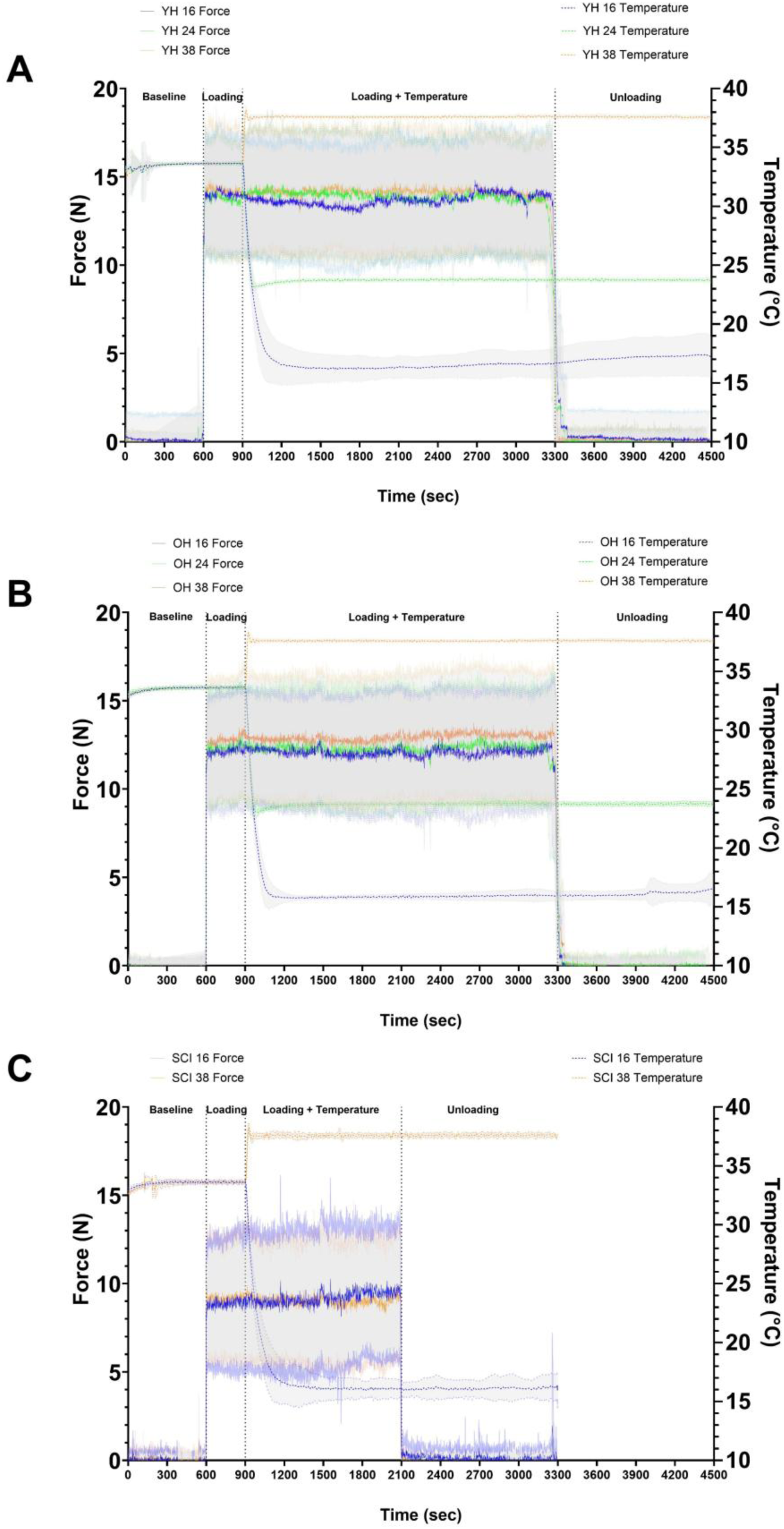
Force and temperature data recorded at the thermo-mechanical indenter probe interface during the protocol. A: data from young healthy group (YH), B: data from older healthy group (OH) and C: data from the spinal cord injury group (SCI). Mean data shown, with SD shaded.

Feasibility and acceptability of our protocol were achieved with minimal issues in the healthy cohorts; however, the significant time commitment required to attend 3 separate sessions on different days, alongside the clinical challenges associated with mobility and day-today variations in health status, meant that this protocol provided significant barriers to accessibility for the SCI cohort. Following consultations with clinical teams at our HRA-approved participants identification centres (i.e. Stoke Mandeville Hospital and Royal National Orthopaedic Hospital, UK) and SCI patients’ representatives, we proceeded with a modification to our protocol to accommodate the needs of SCI participants. Specifically, we a) reduced the protocol’s loading period from 45 to 25 minutes; b) removed the mild cooling testing condition (i.e. 24 °C); and c) allowed both temperature conditions to be completed within the same session, introducing a wash-out period of at least 40 minutes. We also adopted a more conservative approach to the level of mechanical loading applied to the sacrum of the SCI group (i.e. ∼45mmHg instead of ∼60mmHg), to limit the risk of damage to already vulnerable skin. As a result of our protocol modification, SCI participants were able to attend a single ∼6-h testing session, which improved recruitment and testing completion for this group significantly.

Average force and temperature data over the 55-min protocol for the SCI cohort are presented in **Figure 1C** (note when considering Figure 1: the force measurements was zeroed during the 10-minute baseline period with minimal loading to reflect tare weight).

### Experimental Procedures

Participants attended our laboratory within the Clinical Academic Facility located at Southampton General Hospital (UK) on either 3 separate occasions (i.e. healthy cohorts) or a single visit (SCI group). Laboratory conditions were set to thermo-neutral, i.e. 22-24 °C ambient temperature with 50% relative humidity.

Participants were briefed on procedures, provided their written informed consent, before height and mass were recorded (healthy cohorts only; Model 874; Seca GmbH, Hamburg, Germany). Testing conditions (i.e. 38, 24 and 16 °C) were allocated in a counterbalanced order. Participants laid down in the prone position on a hospital bed, and pillows were used to support the lumbar region by flattening out the sacroiliac joint, reducing any pronounced lordosis that may have impacted on probe contact with the skin. The experimental set up is shown in Figure 2.

**Figure 2:**
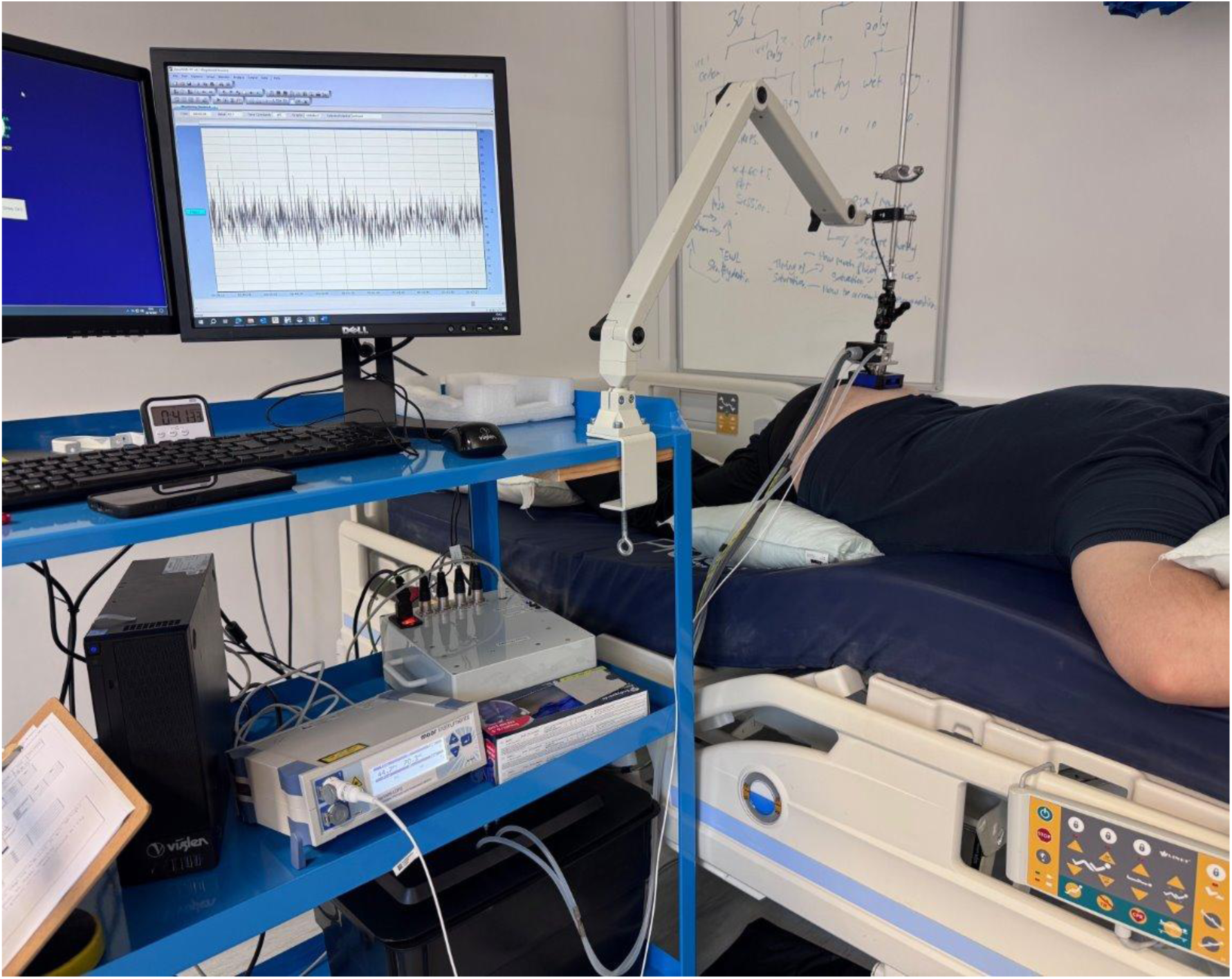
Example of the experimental set up to administer the loading protocol.

Firstly, optical coherence tomography (OCT) was used to capture structural and functional characteristics of the skin of the sacrum. Skin imaging was performed via non-invasive OCT using the VivoSight® handheld device and dynamic OCT processing software (Michelson Diagnostics Ltd., Maidstone, Kent, UK). This technique is non-inferior to punch biopsy for the characterisation of the skin’s epidermis (thickness, stratum corneum hydration, collagen density) and blood perfusion properties (vascular plexus density and diameter) (Adan *et al*., 2022). Detailed methods of this technique can be found in previously published work by our group (Gordon *et al*., 2026). During each experimental session, OCT images were collected in duplicate before loading the sacrum. Following OCT imaging, skin sebum was sampled at the sacrum using Sebutapes (CuDerm, Dallas, TX, USA) for subsequent analysis of inflammatory biomarkers. Biomarker collection and extraction techniques were performed using methods optimised by our laboratory (Soetens *et al*., 2019). Skin sebum sampling was performed before mechanical loading and at the end of the protocol, by application of Sebutapes for 2 minutes using a gloved hand to avoid cross contamination. Sebutapes were then stored at -80 °C prior to analysis using ELISA plates for targeted inflammatory cytokines of interest (e.g. IFN-γ, IL-1β, IL-2, IL-4, IL-6, IL-8, IL-10, IL-12p70, IL-13, TNF-α). The extraction of inflammatory proteins of interest involved both chemical (850 μL PBS + 0.1% Dodecyl maltoside) and physical extraction techniques previously used by our laboratory (Jayabal *et al*., 2023b) using MSD V-Plex plates (Human Proinflammatory Panel 1, Meso-Scale Discovery Maryland, USA). A subset of the sebum samples (see “Data processing and statistical analyses” section) underwent analysis for 71 human cytokines and chemokines (Human Biomarkers Group 1, Meso-Scale Discovery Maryland, USA).

Once pre-loading measures were completed, the thermo-mechanical probe was placed over the sacrum. Before starting the formal loading protocol, pressure distribution at the sacrum was mapped using a pressure mapping device (ForeSiteSS, XSensor, Canada). This was used to select an individualised weight mass required to load the probe and elicit a consistent and uniformly distributed target pressure for each participant (i.e. ∼60mmHg in the healthy cohort and ∼45mmHg for the SCI group). At this point, the protocol commenced and proceed through the 4 phases described in the “Experimental Design” section (i.e. baseline period with minimal loading, loading period with no temperature manipulation, loading period with temperature manipulation, and unloading phase with minimal loading).

Alongside force and temperature sensors, an optic fibre connected to a Laser Doppler Flowmetry (LDF) data acquisition device (Moor Instruments, Axminster, Devon UK) was integrated within the probe and sat flushed at its centre. This allowed continuous measurement of SkBF at the mechanically loaded sacrum during all loading and unloading phases. SkBF was continuously sampled at 40 Hz. Individual SkBF recorded during the 10-minute baseline were used to normalise variables of interest and account for inter-individual variability in resting SkBF levels (see “Data processing and statistical analyses” section).

Perceptual assessment of the participants’ local thermal sensation, comfort and acceptance were also assessed i) at the start and end of the baseline period with minimal loading; ii) at the end of the loading period with no temperature manipulation; and iii) every 10-min during the loading and unloading periods with temperature manipulation; using Likert scales based on recommendations from Schweiker et al., (2020). Thermal sensation consisted of a 7-point scale from 1 (cold) to 7 (hot) and thermal comfort consisted of a 5-point scale ranging from 1 (comfortable) to 5 (extremely uncomfortable). Thermal acceptance consisted of a 4-point scale ranging from 1 (clearly acceptable) to 4 (clearly unacceptable).

Within 2 minutes of completion of the full protocol, post-protocol OCT scanning and skin sebum sampling were performed as described above. Participants were then de-instrumented, debriefed, and left the laboratory.

### Data processing and statistical analyses

Owing to the differences in the experimental loading protocols between the healthy cohorts and SCI group (i.e. shorter duration and 1-less temperature level), we took a pragmatic approach to separate data analyses (i.e. healthy cohorts only vs. SCI group only) for specific datasets (e.g. SkBF vs. inflammatory biomarkers), in order to address our primary hypotheses (i.e. temperature modulation of responses to loading) and secondary hypotheses (i.e. interaction of temperature-dependent responses with age and clinical status). For example, SkBF data were analysed separately for the healthy and SCI cohorts, as evidence indicates that the duration of loading-induced ischemia can modify post-occlusive reactive hyperaemia (Liao *et al*., 2013). Conversely, inflammatory biomarker analyses were grouped across all 3 groups, as evidence on the time-course of acute skin inflammation in response to varying durations of mechanical loading remain limited (Shutova & Boehncke, 2022; Feldt *et al*., 2024).

As previously noted, the loading and temperature profiles of the thermo-mechanical probe were recorded for all participants across all testing. These were analysed separately for healthy and SCI groups for the independent and interactive effects of phase, temperature, and group (healthy only) by means of 3-way mixed method and 2-way repeated measures ANOVA, respectively. This analysis interrogated the consistency of our thermo-mechanical stimuli across tests and participants, and against target loads and temperatures. Statistical analysis was performed using IBM SPSS Statistics v.30 (IBM, Armonk, NY, USA).

### Primary experimental outcomes

As noted in the methods, SkBF responses were selected as primary outcomes, with post-occlusive reactive hyperaemia representing the primary variable informing sample size calculations.

Regarding SkBF data from the healthy cohorts, a low pass Butterworth filter was applied to all recordings to filter respiration and cardiac frequencies, before the data was down sampled to a frequency to 1 Hz. At this point, total area under the curve (AUC) was calculated for each phase of the loading protocol (i.e. baseline period with minimal loading, loading period with no temperature manipulation, loading period with temperature manipulation, and unloading phase with minimal loading) for each participant and for each temperature condition. AUC data for each of the 4 phases were assessed for normality, and depending on data distribution, were analysed separately for the independent and interactive effects of temperature condition (3 levels: 16 °C, 24 °C or 38 °C) and participant age (2 levels: younger or older) using a 2-way mixed model ANOVA or Friedman test. Post-hoc analyses based on significant main effects were performed with Bonferroni-adjusted pairwise comparisons. Furthermore, several variables indicative of specific microvascular responses were calculated at selected time points over the protocol and normalised over the last minute of the baseline period, to account for inter-individual variability. These variables are summarised in **Table 2**, alongside their calculations and interpretation.

**Table 2:** Summary of calculated variables of interest for the skin blood flow (SkBF) data during the protocol.

| SkBF parameter | Calculation | Interpretation |
| --- | --- | --- |
| **1. Change in SkBF during loading only phase** | $\frac{\text{Mean SkBF during loading only phase} \times 100}{\text{Mean SkBF during baseline period}}$ | % change in SkBF from individual baseline resulting from mechanical loading only |
| **2. Change in SkBF during loading period with temperature manipulation (first 5 minutes)** | $\frac{\text{Mean SkBF during loading period with temperature manipulation (first 5 minutes)} \times 100}{\text{Mean SkBF during baseline period}}$ | % change in SkBF from individual baseline resulting from mechanical loading with temperature manipulation at the start of the combined loading and temperature |
| **3. Change in SkBF during loading period with temperature manipulation (last 5 minutes)** | $\frac{\text{Mean SkBF during loading period with temperature manipulation (last 5 minutes)} \times 100}{\text{Mean SkBF during baseline period}}$ | % change in SkBF from individual baseline resulting from mechanical loading with temperature manipulation at the end of the combined loading and temperature |
| **4. Peak post-occlusive reactive hyperaemia** | $\frac{\text{Peak SkBF value during first 3 minutes of unloading} \times 100}{\text{Mean SkBF during baseline period}}$ | % change in SkBF from individual baseline resulting from mechanical unloading with ongoing temperature manipulation |
| **5. Estimated post-occlusive thermal hyperaemia component** | $\frac{\text{Mean SkBF value during last 3 minutes of unloading} \times 100}{\text{Mean SkBF during baseline period}}$ | % change in SkBF from individual baseline at the end of the mechanical unloading with ongoing temperature manipulation to estimate likely thermal hyperaemia component of the post-occlusive response |

The five variables presented in Table 2 were assessed for normality, and depending on data distribution, were analysed separately for the independent and interactive effects of temperature condition (3 levels: 16 °C, 24 °C or 38 °C) and participant age (2 levels: younger or older) using a 2-way mixed model ANOVA or Friedman test. Post-hoc analyses based on significant main effects were performed with Bonferroni-adjusted pairwise comparisons.

**Figure 3** provides a visual example of group-level SkBF responses in the young cohort with annotated phases and SkBF variables described in the preceding sections.

**Figure 3:**
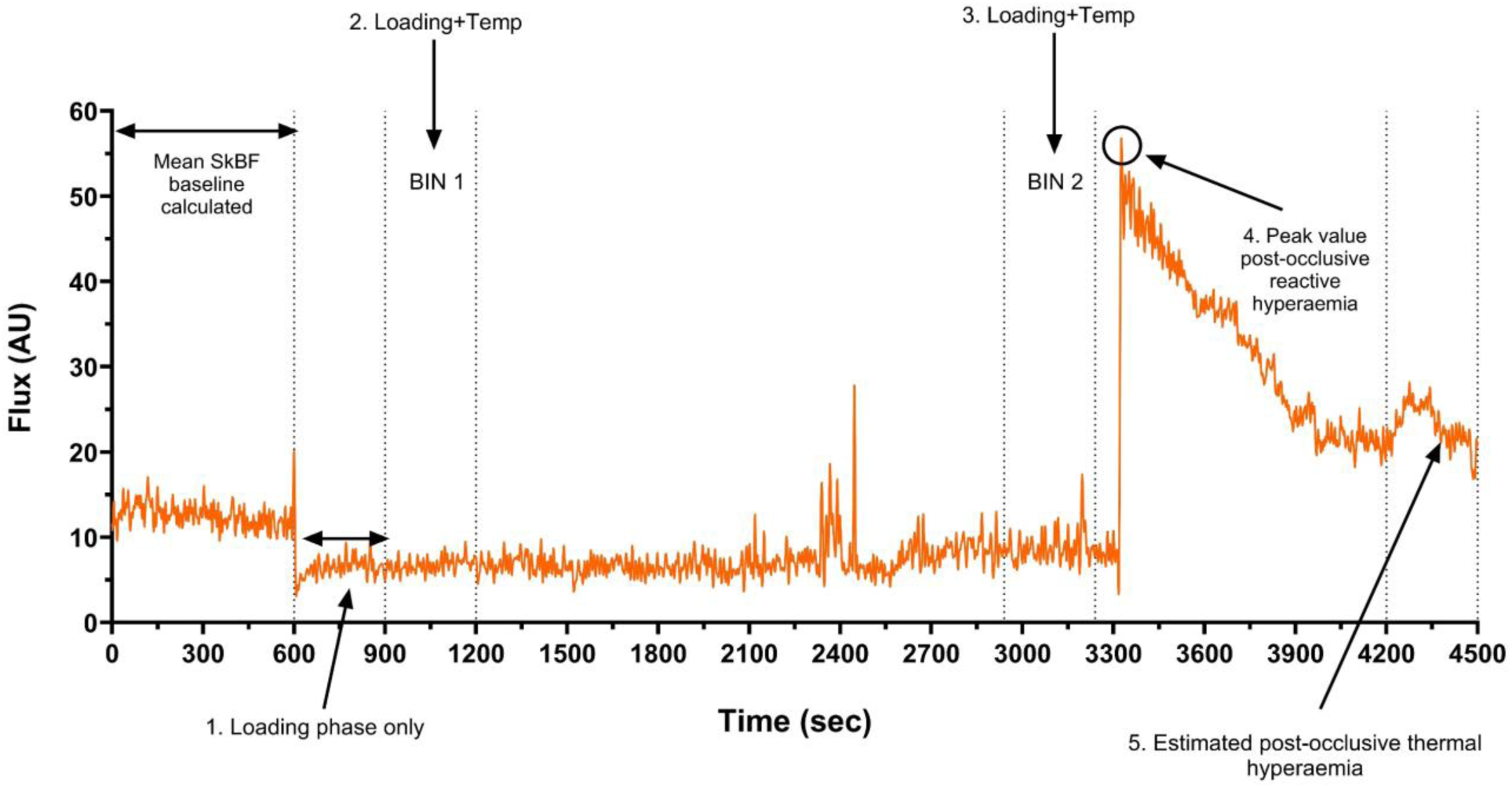
Example of the loading protocol at the sacrum (n=1) in the 38°C with annotated analysis phases of interest. Annotated analysis phases of interest refer to numbered phases and calculations detailed in Table 2.

The same SkBF analyses above were applied to the SCI cohort data, with the only difference being that the SCI dataset was analysed for between-conditions effects only (i.e. 2 levels: 16 °C vs. 38 °C) using paired samples t-tests or Wilcoxon signed-ranks tests.

### Secondary experimental outcomes

Regarding OCT imaging data, scans collected at baseline were processed and analysed using methods previously described (Gordon *et al*., 2026). Variables analysed included optical attenuation coefficient (OAC), skin roughness, vascular density as function of depth, and plexus depth. Healthy cohorts’ data have been published elsewhere (Gordon *et al*., 2026) and were combined with SCI participants’ data (previously unreported) to allow a 3-groups comparison.

Regarding biomarker data, we first analysed skin sebum for a predefined panel of 10 proinflammatory cytokines (IFN-γ, IL-1β, IL-2, IL-4, IL-6, IL-8, IL-10, IL-12p70, IL-13, TNF-α). Following on this analysis, and that from collaborators (Feldt *et al*., 2024), we then followed with an untargeted proteomics approach (71 analytes of interest) in a subset of samples from all 3 experimental groups, to broaden the scope of the biomarker analysis. For all assays, the light intensity for standard concentrations were measured through a Meso Quikplex SQ 120 instrument and the concentration of the samples quantified based on the standard curve for each analyte. For the proteomic analysis, multivariate data analysis was performed using SIMCA software (version 18.0; Sartorius Stedim Biotech, Umeå, Sweden). Proteins below the limit of detection in more than 50% of the study population were excluded from further analysis. A principal component analysis (PCA) was performed to check for outliers using score plots and Hotelling’s T2 and distance to model in X-space. An orthogonal partial least square discriminant analysis (OPLS-DA) was used to determine which cytokines/chemokines were important for class differences in temperature conditions (16 °C and 38 °C) before and after mechanical loading at the sacrum. The variable influence of projection (VIP) was used to determine the importance of each variable. P(corr) represents the loading of each variable scaled as a correlation coefficient. VIP > 1.0 and p(corr) > 0.3 were considered as significant.

Thermal sensation, comfort, and acceptability scores recorded at the end of loading, e.g. 55 mins for healthy groups and 35 mins for SCI, were used to characterise perceptions to loading and temperature combined. The percentage of participants that reported thermal acceptability as ‘clearly acceptable’ was also calculated for this time point.

## Results

### Thermo-mechanical probe loading and temperature profiles

In the healthy cohorts, our analyses indicated that the protocol achieved the target levels of 60-mmHg mechanical loading (**Fig. 1A &B**), evidenced by a main effect of phase (F(2, 80) = 825.86, p < 0.001, η^2^ = 0.954), a 13.17 N increase in force from baseline to loading phases (95% CI: 12.02 to 14.31, p < 0.001), and a return to baseline levels of force during unloading (loading versus unloading mean difference= -13.09 N, 95% CI: -14.22 to -11.96, p < 0.001; **Figures 1A and 1B**). When converted to pressure values (note the probe’ 25cm^2^-surface), the force levels recorded during loading corresponded to ∼57mmHg (i.e. ∼39.5mmHg recorded during loading in addition to ∼17.5mmHg from tare weight). These loading levels were consistently applied between participants groups (i.e. no effect of group; F(1,40) = 2.85, p = 0.099, η^2^ = 0.067), and across temperature conditions (no effect of temperature; F(2,80) = 0.94, p =0.385, η^2^ = 0.023). Similarly, the SCI data indicated that the protocol achieved the target levels of 45-mmHg mechanical loading (**Fig. 1C**), evidenced by a main effect of phase (F(2,18) = 69.4, p < 0.001, η^2^ = 0.885), a 9.1 N increase in force from baseline to loading phases (95% CI: 5.9 to 12.3, p < 0.001), and a return to baseline levels of force during unloading (loading versus unloading mean difference= -9.1 N, 95% CI: - 12.3 -to -5.9, p < 0.001). When converted to pressure values (note the probe’ 25cm^2^-surface), the force levels recorded during loading corresponded to ∼45mmHg (i.e. ∼27.3mmHg recorded during loading in addition to ∼17.5mmHg from tare weight). These loading levels were consistently applied between temperature conditions (no effect of temperature; (F(1, 9) = 0.380, p = 0.553, η^2^ = 0.041).

Regarding the probe’s temperature profiles, our analyses of data in the healthy groups indicated that the protocol achieved target temperature levels across all conditions (**Fig. 1A &B**), as evidenced by a main effect of condition (F(2,80) = 15555.01, p <0.001, η^2^ = 0.954), phase (F(1, 40) = 25113.5, p < 0.001, η^2^ = 0.997), and an interaction between condition and phase (F(2,80) = 15400.2, p <0.001, η^2^ = 0.997). Post-hoc comparisons showed no difference between conditions during baseline (mean probe temperature younger: 33.5 ± 0.2 °C; older: 33.5 ± 0.0; all p > 0.05), and clear differences during all loading and unloading phases (all p<0.001), with average probe temperatures being consistently maintained at either 16.7±1.0 °C, 23.8±0.0 °C and 37.6 ± 0.0 °C depending on the testing condition. These temperature profiles were consistently achieved between participants groups (i.e. no effect of group; F(1, 40) = 3.53, p = 0.068, η^2^ = 0.081). Similarly, the SCI data indicated that the protocol achieved the target levels of temperature, with main effects of condition (F(1, 9) = 4060.1, p < 0.001, η^2^ = 0.998) phase (F(1, 9) = 1676.8, p < 0.001, η^2^ = 0.995), and condition by phase interaction (F(1, 9) = 3585.7, p < 0.001, η^2^ = 0.997). Post-hoc comparisons showed no difference between conditions during baseline (mean probe temperature= 33.5 ± 0.1 °C; p = 0.257), and clear differences during all loading and unloading phases (all p<0.001), with average probe temperatures being consistently maintained at either 16.7 ± 1.0 °C and 37.6 ± 0.0 °C depending on the testing condition.

### Skin blood flow

**Table 3** summarises SkBF responses at the sacrum during the loading and unloading protocol for all groups.

**Table 3:** Calculated skin blood flow (SkBF) variables during the protocol at the sacrum for all participant groups. Data presented as mean ± SD. Difference from 38°C indicated as * = p<0.05; ** = p<0.01 and *** = p<0.001. ^###^ indicates difference from 24 °C at p<0.001 level. AUC: area under the curve.

| Age Group | Young Healthy (n = 23) |  |  | Old Healthy (n = 19) |  |  | Spinal Cord Injury (n = 10) |  |
| --- | --- | --- | --- | --- | --- | --- | --- | --- |
|  | 38°C | 24°C | 16°C | 38°C | 24°C | 16°C | 38°C | 16°C |
| AUC Baseline | 7411 ± 2531 | 8005 ± 4231 | 8251 ± 3731 | 6487 ± 2162 | 8342 ± 4611 | 7522 ± 3541 | 6273 ± 1834 | 7701 ± 2631* |
| AUC loading only | 3137 ± 1522 | 3017 ± 1000 | 3144 ± 1542 | 2468 ± 1214 | 3404 ± 2309 | 2630 ± 995 | 3056 ± 1041 | 3789 ± 2165 |
| AUC loading with temperature modulation | 28567 ± 18272 | 15573 ± 6476*** | 13853 ± 7799*** | 29588 ± 21851 | 17272 ± 13937*** | 10773 ± 10335*** | 17275 ± 6245 | 7850 ± 4650*** |
| AUC unloading with temperature modulation | 43107 ± 21324 | 11975 ± 10322*** | 16675 ± 12343*** | 44959 ± 20884 | 9832 ± 7018*** | 13055 ± 10486*** | 35537 ± 13967 | 10685 ± 7816*** |
| Normalised effect of loading only (%) | -18.1 ± 24.1 | -15.7 ± 27 | -23.0 ± 23.5 | -26.4 ± 21.2 | -21.0 ± 18.9 | -22.33 ± 21.7 | -4.4 ± 25.5 | -5.5 ± 26.9 |
| Normalised effect of loading and temperature modulation BIN 1 (%) | -6.0 ± 43.0 | -46.1 ± 21.7*** | -62.8 ± 14.2***### | -11.0 ± 38.7 | -53.5 ± 18.6*** | -67.2 ± 15.7***### | 36.9 ± 60.1 | -56.8 ± 18.3*** |
| Normalised effect of loading and temperature modulation BIN 2 (%) | 9.6 ± 65.7 | -47.3 ± 19.7*** | -57.2 ± 22.6*** | 47.7 ± 105.1 | -51.4 ± 17.8*** | -56.4 ± 46.5*** | 65.2 ± 82.3 | -52.8 ± 18.8*** |
| Normalised peak hyperaemia (%) | 388.4 ± 282.1 | 76.6 ± 94.7*** | 56.7 ± 93.9*** | 377.9 ± 219.8 | 61.1 ± 101.8*** | 50.9 ± 71.6*** | 207.4 ± 151.5 | 1.7 ± 39.1** |
| Estimated post-occlusive thermal hyperaemia component (%) | 141.9 ± 97.3 | -25.9 ± 67.5*** | -0.1 ± 97.7*** | 200 ± 111 | -48.1 ± 16.6*** | -23.4 ± 58.0*** | 219.5 ± 155.6 | -32.8 ± 51.6** |

In the healthy cohorts, SkBF AUC at baseline did not differ between temperature conditions (F(2,80) = 2.179, p = 0.130, η^2^ = 0.052) nor groups (F(1,40 = 0.264, p = 0.610, η^2^ = 0.007) (**Fig. 4A & B**). Upon application of loading only, SkBF decreased on average by 21% (i.e. see “Normalised effect of loading only (%)” – **Tab. 3**), with no differences in SkBF AUC between temperature conditions (F(2,78) = 1.656, p = 0.198, η^2^ = 0.041) nor groups (F(1, 39) = 0.491, p = 0.487, η^2^ = 0.012) (**Fig. 4A & B**). Upon combination of loading with temperature modulation, further changes in SkBF occurred (**Fig. 4A & B**). Specifically, SkBF decreased further during this phase to reach average values ∼53% below baseline during the 24 and 16°C conditions (i.e. see “Normalised effect of loading and temperature modulation BIN 2 (%)” – **Tab. 3**). Conversely, this decrease was not observed during the 38°C condition. In support of this observation, analyses of SkBF AUC during loading with temperature modulation indicated a main effect of temperature (F(2,78) = 22.19, p <0.001, η^2^ = 0.363), with a two-fold difference between the 38 °C condition and the 16 °C (mean difference= 16764 AUs, 95% CI: 23857 to 9672, p < 0.001) and 24 °C condition (mean difference: 12655 AUs, 95% CI: 20354 to 4965, p < 0.001). These temperature-dependent differences in SkBF AUC during loading with temperature modulation occurred similarly in both age groups (i.e. no main effect of group; F(1,39) =, p = 0.970, η^2^ = 0.000) (**Fig. 4A & B**). Upon removal of the loading, normalised peak hyperaemia responses varied greatly as a function of temperature condition (main effect; F(2,80) = 74.92, p < 0.001, η^2^ = 0.652) but not age group (F(1,40) = 0.08, p = 0.779, η^2^ = 0.002). Specifically, SkBF increased above baseline levels within the first 3 minutes of unloading by an average of 382% in the 38 °C, 49% in the 24 °C, and 53% in the 16 °C condition (**Tab. 3**). Post-hoc comparisons indicated that, relative to the 38°C condition, the peak hyperaemic responses were attenuated by 314.3% in the 24 °C (95% CI: 229.3 to 399.3, p < 0.001) and by 329.4% in the 16 °C condition (95% CI: 235.7 to 423.1, p < 0.001). In support of this observation, analyses of SkBF AUC during unloading with temperature modulation indicated a main effect of temperature (F(2,78) = 77.4, p < 0.001, η^2^ = 0.665) but not age group (F(1,39) = 0.152, p = 0.669, η^2^ = 0.004), with a three-fold difference between the 38 °C condition and the 16 °C (mean difference= 29168 AUs, 95% CI: 21193 to 37143, p < 0.001) and the 24°C condition (mean difference= 33129 AUs, 95% CI: 24486 to 41773, p < 0.001). Peak hyperaemic responses decreased over the unloading period with temperature modulation, such that they reached average levels ∼24% below baseline during the last 3 minutes of the 24 °C and 16 °C conditions, and ∼170% above baseline at the end of the 38 °C condition (**Tab. 3**). This is supported by a main effect of temperature (F(2,80) = 98.30, p < 0.001, η^2^ = 0.711) and an interaction effect (F(2,80 = 4.6, p = 0.027, η^2^ = 0.094), for the estimated effect of thermal hyperaemia. Yet, there were no differences detected between age groups in relative blood flow responses in the final 3 minutes in the 16 °C (23.2%, 95% CI: -28.3 to 74.8, p = 0.368, 24 °C (22.2 %, 95% CI: -9.9 to 54.4, p = 0.170) or 38 °C temperature conditions (58.1%, 95% CI: -6.8 to 123.1, p = 0.078).

**Figure 4:**
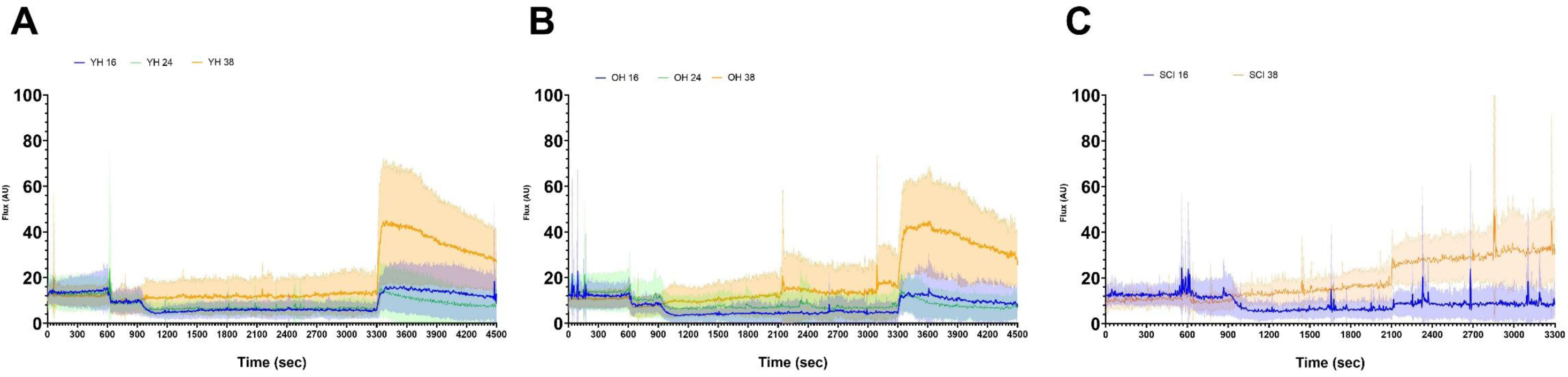
Mean skin blood flow traces for each temperature condition: 16 °C, 24°C and 38 °C. Shaded area represents standard deviation. A: Young healthy group (YH; N= 23). B: older healthy group (OH; N= 19) and C: spinal cord injury (SCI; N= 10).

In the SCI cohort, SkBF AUC at baseline was significantly different between temperature conditions (t(9) = 2.396; p = 0.040; 1428 AU·s, 95% CI: 80 to 2777). With the application of loading only, SkBF decreased similarly (∼5%) in both temperature conditions with no differences in SkBF loading AUC between 16 and 38 °C (t(9) = 1.677; p = 0.128; 733 AU·s, 95% CI: -2.56 to 1722). Upon addition of temperature to the load, further changes to SkBF occurred, such that it decreased further to 53% below baseline in the 16°C condition; or it increased to 65% above baseline in the 38°C condition. This is supported by the statistically significantly larger SkBF AUC calculated during loading and temperature modulation in the 38°C compared to 16°C (t(9) = 6.642; p < 0.001; 9425 AU·s, 95% CI: 6215 to 12636). Upon the removal of the loading, normalised post-occlusive peak hyperaemia responses differed by temperature (t(9) = 4.141; p = 0.003; mean difference 16 vs. 38 °C= 206%, 95% CI: 93.3 to 318). Specifically, within the first 3 minutes of unloading, SkBF increased above baseline by an average of 207% during the 38°C condition, and by 2% in the 16°C conditions. This observation in corroborated by the distinct differences between temperature conditions for analyses of SkBF AUC during unloading with temperature modulation (t(9) = 4.726; p = 0.001; mean difference 16 vs. 38 °C= 24853 AU·s, 95% CI: 12957 to 36749). Post-occlusive peak hyperaemic responses varied over time during the unloading period with temperature modulation, such that they reached average levels of 33% below baseline during the last 3 minutes of the 16°C condition, and 220% above baseline during the last 3 minutes of the 38°C condition (**Tab. 3**; t(9) = 4.511; p = 0.001; mean difference 16 vs. 38 °C= 252%, 95% CI: 126 to 379).

### Skin imaging

Skin epidermal and microvascular properties for the three groups are summarised in **Table 4**. Assessment of skin properties at the sacrum at baseline did not show any differences between young, older and SCI participants for optical attenuation coefficient (OAC; F(2,48) = 1.757, p = 0.184, η^2^ = 0.071), dermal brightness (F(2,48) = 1.215, p = 0.306, η^2^ = 0.051), skin roughness (F(2,49) = 0.163, p = 0.850, η^2^ = 0.007), nor plexus depth (F(2,50) = 2.925, p = 0.063, η^2^ = 0.109).

**Table 4:** Skin epidermal (i.e. optical attenuation coefficient, dermal brightness, skin roughness) and microvascular properties (plexus depth) at the sacrum for the younger, older and spinal cord injured cohort. Data are presented as mean±SD and actual sample size available for analyses (N) is reported for each parameter.

| Skin Parameter | Younger | Older | Spinal Cord Injured |
| --- | --- | --- | --- |
| Optical Attenuation Coefficient (mm<sup>-1</sup>) | 2.85±0.23 | 2.92±0.31 | 2.73±0.20 |
| N | 22 | 17 | 10 |
| Dermal Brightness (%) | 93.6±4.4 | 93.0±4.64 | 90.8±1.83 |
| N | 22 | 17 | 10 |
| Rq (μm) | 20.4±4.3 | 19.6±4.7 | 20.0±4.4 |
| N | 22 | 18 | 10 |
| Plexus Depth (μm) | 269.2±30.1 | 276.5±43.4 | 302.2±32.3 |
| N | 22 | 19 | 10 |

Vascular density profiles of the sacrum at baseline for the three groups are shown in **Figure 5**. Vascular density varied as a function of depth (0.05 – 1.0 mm) and group, as indicated by a statistically significant main effect of depth (p < 0.001), group (p < 0.001) and interaction of depth by group (p < 0.014). Specifically, post-hoc comparisons showed that vascular density was greater in the SCI cohort (range: +0.41 to +3.36%; all p < 0.05) at all depths, except for the 0.20 – 0.35mm range, and the 1.0mm depth (**Fig. 5**).

**Figure 5:**
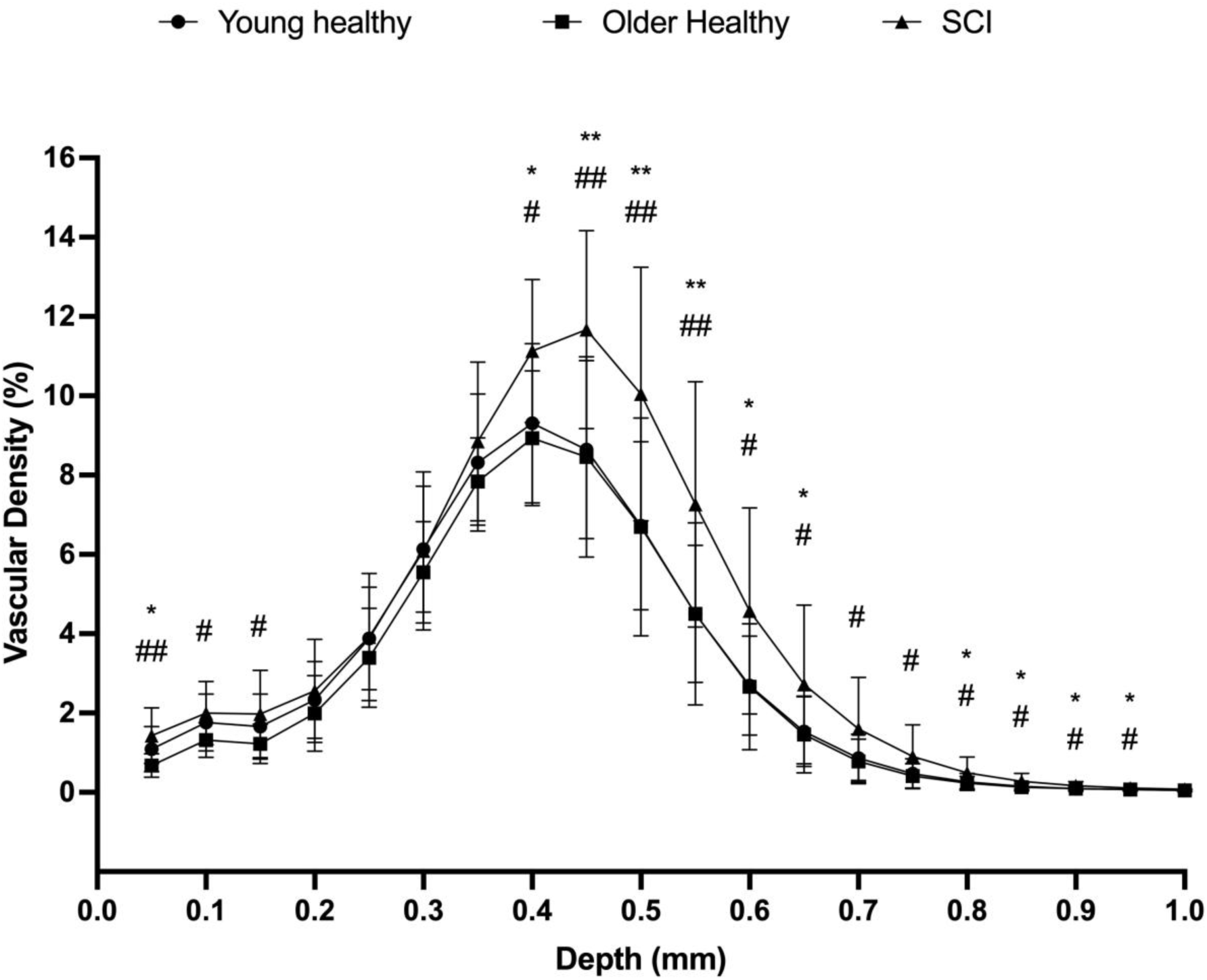
Vascular density profiles of the sacrum for younger healthy (n = 22), older (n = 19) and spinal cord injured (SCI; n =10) groups plotted against depth. Data are mean±SD. * Denotes significant difference in the SCI group (p<0.05) from the young and # denotes difference (p<0.05) from older group. ** Denotes significant difference in the SCI group (p<0.01) from the young and ## denotes difference (p<0.01) from older group.

### Inflammatory response to loading

#### Targeted pro-inffammatory panel

The results of the analysis of targeted pro-inflammatory cytokines are summarised in **Table 5**. For all pro-inflammatory cytokines measured in the healthy cohorts (N=20), a statistically significant main effect of time was detected (all p < 0.005; **Tab. 5**), whereby concentrations of all cytokines decreased following mechanical loading and unloading (range of decrease= -0.39 to -5.33 pg/mL). These responses did not differ between age groups nor temperature condition (i.e. no statistically significant effects of age, temperature condition, or interaction; all p > 0.05).

**Table 5:**
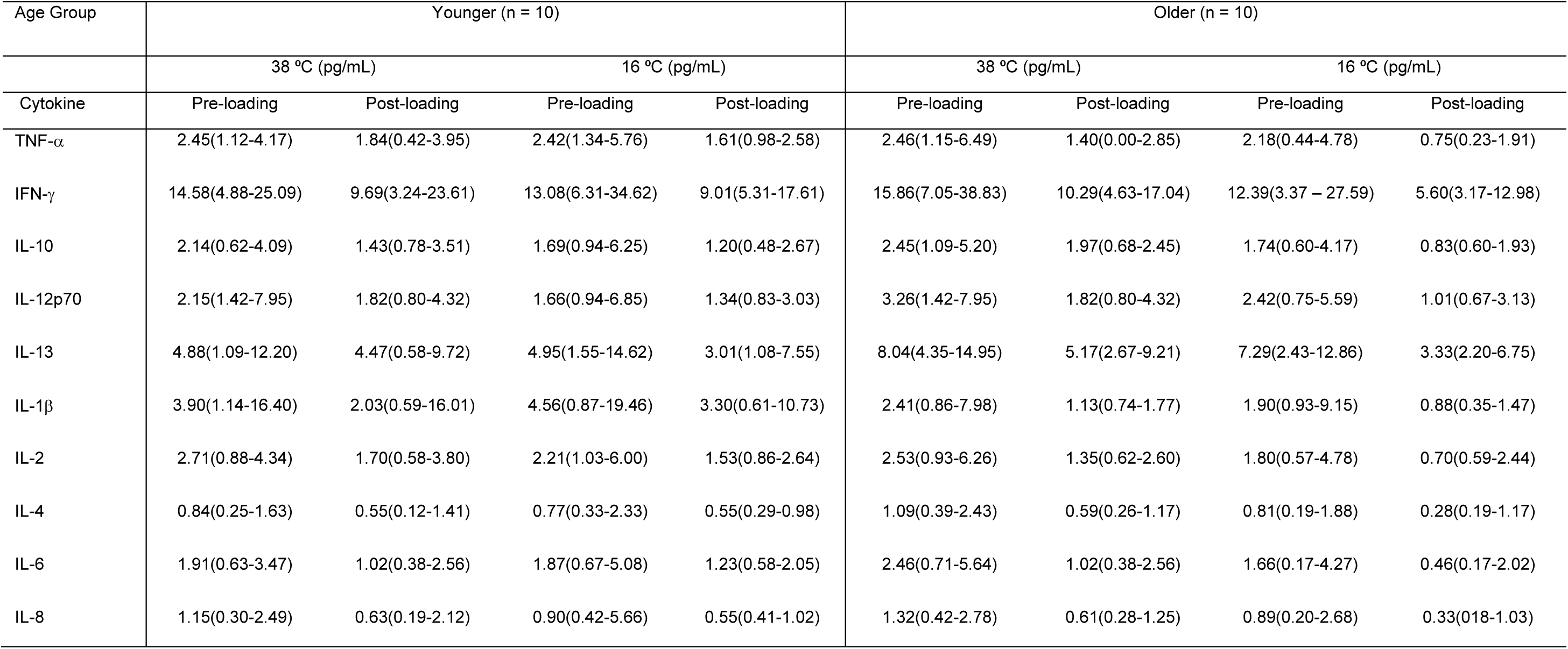
Concentration of pro-inflammatory cytokines measured in skin sebum pre- and post-mechanical loading in 16 °C and 38 °C. These data are from a subset of the participants, n=10 in both healthy cohorts. Data presented as median(range).

#### Untargeted proteomics approach

Untargeted proteomics analyses were completed for a further subset of samples from the 3 cohorts (young N= 6, old N= 6, and SCI N= 8). Multivariate statistical analysis in the 29 proteins of interest that were above the limit of detection were used to investigate the differences in protein response before and after the application of mechanical load and by temperature condition. Analysis using an OPLS-DA showed separation by timepoint only, with sensitivity (R^2^ = 0.395), predictivity (Q^2^ = 0.164) with a statistically significant CV-ANOVA (p < 0.05), whereby a generalised decrease in cytokines was identified post loading. Following OPLS-DA analysis, 10 proteins were identified to contribute most to the separation (YKL-40, IL-1RA, IL-1α, MCP-2, Fractalkine, IFN-α2α, TSLP, IL-1β, IL-21 and IL-31) using the threshold VIP > 1 and p(corr)>0.3. Concentrations of these 10 proteins of interest are summarised in **Table 6**.

**Table 6:** Concentration of ten proteins identified to drive separation in PCA analysis measured in skin sebum pre- and post-mechanical loading in 16 °C and 38 °C. These data are from a subset of the participants, younger n=6, older n=6 and spinal cord injured (n=8. Data presented as median(range).

| Age Group | Younger |  |  |  | Older |  |  |  | Spinal Cord Injured |  |  |  |
| --- | --- | --- | --- | --- | --- | --- | --- | --- | --- | --- | --- | --- |
|  | 38 °C (pg/mL) |  | 16 °C (pg/mL) |  | 38 °C (pg/mL) |  | 16 °C (pg/mL) |  | 38 °C (pg/mL) |  | 16 °C (pg/mL) |  |
| Cytokine | Pre-loading | Post-loading | Pre-loading | Post-loading | Pre-loading | Post-loading | Pre-loading | Post-loading | Pre-loading | Post-loading | Pre-loading | Post-loading |
| YKL-40 | 248.5(2064.4) | 87.4(498.0) | 332.4(1545.7) | 180.1(979.2) | 398.8(3329.6) | 92.4(1326.9) | 693.4(2293.4) | 142.6(2832.9) | 398.1(1061.8) | 86.2(516.4) | 504.2(1140.2) | 229.3(650.0) |
| IL-1RA | 468.0(1243.1) | 66.1(346.5) | 486.6(756.8) | 103.8(466.5) | 182.2(478.4) | 33.3(298.6) | 304.6(1120.7) | 136.8(278.0) | 287.2(4478.8) | 58.2(420.2) | 287.8(512.3) | 92.7(1012.3) |
| IL-1α | 338.3(1770.3) | 93.6(1249.2) | 258.4(1476.5) | 154.1(824.4) | 567.3(3713.7) | 222.8(1405.1) | 918.7(2007.2) | 215.0(3032.2) | 805.9(2942.0) | 414.9(1033.7) | 1173.0(2066.2) | 345.8(2318.4) |
| MCP-2 | 0.03(0.07) | 0.04(0.13) | 0.04(0.08) | 0.06(0.09) | 0.02(0.99) | 0.04(0.07) | 0.02(0.04) | 0.02(0.03) | 0.02(0.03) | 0.02(0.05) | 0.02(0.03) | 0.03(0.04) |
| Fractalkine | 63.4(172.4) | 58.7(99.1) | 64.2(162.4) | 21.8(57.4) | 89.4(1555.1) | 46.8(95.1) | 134.6(164.5) | 20.9(74.2) | 22.1(82.3) | 22.1(65.5) | 34.2(141.2) | 25.7(397.3) |
| IFN-α2a | 0.2(3.3) | 0.2(2.0) | 0.2(0.9) | 0.2(0.2) | 0.2(28.2) | 0.2(0.9) | 0.4(2.0) | 0.2(2.2) | 0.5(6.0) | 0.2(0.9) | 0.3(1.2) | 0.2(2.1) |
| TSLP | 0.2(0.3) | 0.2(1.2) | 0.2(0.6) | 0.3(0.3) | 0.4(3.6) | 0.5(1.6) | 0.6(1.1) | 0.2(0.6) | 0.2(0.3) | 0.2(0.5) | 0.2(1.0) | 0.4(0.9) |
| IL-1β | 2.0(7.6) | 1.0(3.9) | 1.3(2.7) | 0.7(1.2) | 1.2(9.0) | 1.1(2.3) | 2.5(13.8) | 1.4(4.9) | 1.2(2.6) | 0.8(2.2) | 1.7(3.3) | 0.7(4.3) |
| IL-21 | 2.8(3.0) | 2.9(8.6) | 5.6(5.9) | 3.8(5.3) | 5.7(58.9) | 5.7(6.6) | 3.2(8.6) | 2.9(6.2) | 3.1(6.5) | 3.2(4.4) | 2.6(2.3) | 4.3(6.2) |
| IL-31 | 9.5(11.7) | 15.3(10.2) | 13.3(12.6) | 14.6(3.8) | 13.2(56.4) | 14.0(17.3) | 9.6(14.7) | 10.3(17.4) | 10.9(7.0) | 14.0(10.4) | 12.1(12.8) | 12.1(14.8) |

### Perceptual responses

At the point of unloading (55min), 70% and 78% of healthy young participants reported ratings of ‘thermally acceptable’ for the 16- and 24-°C conditions, respectively. At the same time point, 95% and 89% of healthy older participants, reported ratings of ‘thermally acceptable’ for the 16- and 24-°C conditions, respectively.

Seven of the ten SCI participants had residual sensation at the sacrum. 86% of this group (6/7) reported ratings of “thermally acceptable” This translated into 6 of 7 participants rated the cooling (16 °C) as ‘thermally acceptable’ for the 16°C condition at the point of unloading (35min).

Regarding thermal sensation, the median score at the point of unloading (55min) in the healthy cohorts was 3 in both the younger and older cohorts, reporting thermal sensation as ‘Slightly Cool’. Similarly, in our SCI the median score was also reported as ‘3’ or ‘Slightly Cool at the point of unloading (35 min).

Regarding thermal comfort, the median score at the point of unloading (55min) in the healthy cohorts was 1 or ‘comfortable’ for both the younger and older cohorts. The SCI participants reported similarly with a median score of 1 or ‘comfortable’ at unloading (35 min).

## Discussion

The aim of this study was to investigate how varying levels of localised cooling (i.e. 24 and 16 °C) modulate the microvascular, inflammatory and perceptual responses to mechanical loading and unloading of the skin of the sacrum in young and older adults, and in a sub-cohort of patients at high risk of PU, i.e. those with a SCI. We hypothesised that skin cooling would downregulate both microvascular (e.g. peak post-occlusive reactive hyperaemia) and inflammatory responses to mechanical loading, while potentially upregulating discomfort, in a dose-dependent fashion. We also hypothesised that these responses would be attenuated in aged skin and in the presence of SCI.

Leveraging a large cohort (N=52) of healthy participants and patients, and by combining non-invasive assessments of skin’ microvascular (i.e. SkBF), inflammatory (i.e. skin-sebum biomarkers), structural (i.e. OCT imaging) and perceptual responses to controlled thermo-mechanical insults at the sacrum, our study provides three novel insights on the temperature-modulation of skin tissue viability.

First, milder and more intense skin cooling (i.e. 24 and 16 °C) induced a similar ∼8-fold decrease in peak post-occlusive reactive hyperaemic responses in young and older healthy participants, following a 40-minute loading period with 60mmHg pressure at the sacrum, when compared to a 38 °C control condition. Temperature modulation of SkBF responses to mechanical loading persisted during the 20-min unloading phase, with AUC for SkBF during this phase being 3 times smaller in the 24 and 16 °C cooling conditions than during the 38 °C condition. Similar temperature-dependent differences were observed in SkBF responses in the SCI group, such that peak post-occlusive reactive hyperaemia was largely suppressed at the point of unloading in the 16 °C-(i.e. ∼2% increase in SkBF above baseline) compared to the 38 °C condition (i.e. ∼200% increase in SkBF above baseline). Second, our biomarker analysis indicated that pro-inflammatory cytokines sampled from skin sebum decreased from baseline to the end of the loading and unloading period; yet, this occurred similarly across all temperature conditions and participant groups. Third, most participants (i.e. ≥70%) rated both 24 and 16 °C skin cooling during mechanical loading as thermally acceptable.

Taken together our results indicate that both mild and more pronounced skin cooling are: i) similarly effective in largely suppressing post-occlusive reactive hyperaemia at the sacrum in younger, older and vulnerable skin; and ii) perceptually tolerable by a range of at-risk groups (i.e. older adults and those with a SCI). These findings support the concept that local skin temperature is a potent modulator of the skin microvascular response to mechanical loading; and that cooling the skin during both mechanical loading and unloading could represent a promising PU prevention strategy. The fundamental and applied implications of our findings are discussed in the sections below.

### Skin cooling as a modulator of skin tissue responses to mechanical loading

We believe that the observed attenuation of post-occlusive reactive hyperaemia following unloading in all participants’ cohorts was primarily caused by cooling-mediated changes in local vascular control mechanisms that occurred both during the loading and unloading phases. Indeed, our loading protocol successfully elicited consistent pressure levels at the sacrum across all conditions and groups (see **Fig. 1**), which allowed us to consider the effect of temperature independently of mechanical load.

With respect to the loading phase, it has been previously suggested that cooling the skin during mechanical loading may reduce the tissue’s metabolic demands; this may in turn reduce the extent of skin tissue ischemia and the associated accumulation of vasodilatory metabolites, which are known to directly contribute to the magnitude of post-occlusive hyperaemic responses (Tzen *et al*., 2010; Jan *et al*., 2012; Liao *et al*., 2019; Jan, 2020). For example, in a pilot study involving a small sample (N=10) of 20-40y healthy participants, Tzen et al., have previously observed a ∼50% reduction in post-occlusive hyperaemic responses to 30-min of loading of the sacrum (i.e. ∼60mmHg pressure) between a 25 °C (i.e. cooling) and a ∼28 °C condition (i.e. control) (Tzen *et al*., 2010). This effect was attributed to a likely reduction in metabolic debt during loading with cooling (Hagisawa *et al*., 1994), as evidenced by the observation of a decreased metabolic spectral density during the 25 °C condition (Tzen *et al*., 2010).

Our observations build upon and expands those of Tzen et al. (Tzen *et al*., 2010), as we identified that local skin cooling beyond 25 °C (i.e. down to 16°C) suppressed post-occlusive hyperaemic responses by up to ∼300% in the healthy groups, when compared to a control temperature condition (i.e. 38 °C) that is more likely to represent skin temperature levels occurring under ecologically-valid clinical settings (i.e. consider bedridden patients, who have been reported to experience ∼38 °C skin temperature at the sacrum due to prolonged mechanical loading and thermal insulation of the skin (Yilmaz & Günes, 2019). Importantly, in contrast with the work of Tzen et al. (Tzen *et al*., 2010), our protocol maintained skin cooling throughout the unloading phase, which likely minimised the confounding effects of post-occlusive skin rewarming on local SkBF (Tzen *et al*., 2010; Lee *et al*., 2014).

Considering the above, we believe that our participants’ skin experienced a combination of potent, cooling-mediated vasoconstrictive tone, via activation of sympathetic adrenergic pathways (Johnson *et al*., 2014b), with metabolism-mediated vascular control at the point of unloading. This would have contributed to the large suppression of post-occlusive reactive hyperaemia we observed. The ongoing presence of vasoconstrictor tone throughout our protocol is evidenced by the fact that both 24 and 16 °C conditions resulted in i) a further decrease in SkBF from the loading-only (i.e. SkBF= ∼21% below baseline) to the loading-and-temperature phase (i.e. SkBF= ∼53% below baseline) in the healthy cohorts; ii) the return of SkBF at end of the unloading phase to levels similar to those recorded at baseline (see **Tab. 3**). Furthermore, cooling has been shown to inhibit endothelial nitric oxide synthase enzyme activity, and the resulting post-occlusive, nitric oxide-mediated vasodilation (Johnson & Kellogg, 2018).

An important, yet unexpected finding of this study is that the microvascular responses to mechanical loading were almost identical between the 24 and 16 °C conditions, despite the colder condition approaching the cold-pain threshold (i.e. 15 °C) (Filingeri, 2016). The absence of a dose-response effect would point at the likely presence of “threshold” mechanisms to achieve maximum vasoconstriction, which were likely to be partly mediated by axon reflexes in cold-sensitive Aδ-type cutaneous thermoreceptors (Johnson & Kellogg, 2018). This hypothesis is not entirely speculative. Indeed, Campero et al. have previously reported microneurographic recordings from two human Aδ-type cutaneous thermoreceptors and shown that such fibers responded dynamically to a cooling ramp in the range of 30 °C to 14 °C with a maximum discharge at temperatures ∼27 °C (Campero *et al*., 2009). Based on this evidence, our 24 °C condition would have therefore crossed a relevant cold-receptor threshold, and may have driven the microvasculature close to its constricted maximum, with no additional benefits of cooling to 16°C.

We should also note that, albeit relevant for applications to clinically relevant settings (e.g. bedridden patients), the use of 38 °C temperature for the control condition may have resulted in microvascular responses to unloading that were likely triggered by a combination of both post-occlusive (i.e. ischemia-mediated) and thermal (i.e. warming-mediated) drivers. This may have contributed to inflate the effect size of cooling compared to the control condition. For this reason, we attempted to quantify the relative contributions of thermal and post-occlusive components to the hyperaemic responses by calculating the parameter “Estimated post-occlusive thermal hyperaemia component” (see **Tab.2**), using data from the last 3min of the unloading phase. We believe that this approach provided a conservative estimate of the thermal hyperaemia component (note in **Fig. 4**: SkBF is continuously decreasing during the 38°C condition at the end of the trial). When accounting for the ∼140% elevation in SkBF at the end of the 38 °C condition estimated to arise from thermal hyperaemia (see **Tab. 2**), both 24 and 16 °C skin cooling still resulted in a meaningful five-fold decrease in (post-occlusive) peak hyperaemia.

Healthy ageing has been shown to impact both the structure of the skin and control of the microvasculature, with ageing causing deficits in endothelial nitric oxide availability, reduced vasodilation response due to impaired NOS signalling and increased oxidative stress (Minson, 2010). Despite these changes, age is associated with chronic vasoconstriction in response to localised cooling due to increased local Rho-kinase signalling (Thompson-Torgerson *et al*., 2007). In the present study we did not observe any differences in microvascular response between the younger and older cohorts (**Tab. 3**). A possible explanation is that reduction in metabolic demand and ischaemic burden induced by our cooling levels may have been sufficient to mask these age-related impairments. Additionally, it cannot be excluded that the lack of any disease in our older participants meant that they were less likely to demonstrate age-related declines in skin functionality at the sacrum. The increased vulnerability of aged skin observed clinically is likely the result of a complex interaction between vascular dysfunction, impaired tissue tolerance, co-morbidities and frailty, with incidence of PU development markedly increased in those aged >70 (Jaul *et al*., 2018; Pang *et al*., 2025), i.e. which is older than our aged cohort. OCT scanning in our participants indicated that the structural characteristics of the skin microvasculature were unchanged by age at baseline (Gordon *et al*., 2026); providing additional support for the lack of age-related differences. These findings support the concept that PU risk in older adults is not solely driven by age, but by the cumulative effects of additional risk factors.

When considering our SCI participants, SkBF data indicated that, compared to the 38 °C condition, 16 °C cooling reduced the post-occlusive reactive hyperaemic response to unloading (i.e. normalised peak hyperaemia and AUC unloading with temperature modulation; **Tab. 2**). This finding is consistent with previous studies in SCI patients exposing sacral skin to local cooling (Tzen *et al*., 2010, 2019; Jan *et al*., 2013). Although we cannot directly compare SCI findings with those from our healthy cohorts, due to methodological differences (i.e. duration of loading), the normalised peak hyperaemic response to the 38 °C condition appeared to be weaker in our SCI than in the healthy cohorts (**Tab. 3**). This could be due to several reasons. First, it has been suggested that sensory nerves contribute to the reactive hyperaemic response, which may be attenuated in individuals with SCI (Larkin & Williams, 1993; Lorenzo & Minson, 2007). This has been evidenced by a reduction in neurogenic spectral frequency of reactive hyperaemia in SCI patients undergoing a similar protocol to the one employed in this study (Jan *et al*., 2013). However, our SCI participants demonstrated maintained microvascular regulatory function in response to loading and temperature stimuli (see **Tab. 2** changes in SkBF with phase). It is therefore more likely that the attenuated responses in our SCI cohort were partly due to the reduced levels of mechanical loading that were used in this group (i.e. ∼45mmHg), to minimise additional risks to their skin integrity. This consideration is supported by the observation that SkBF continued to raise in response to the 38 °C during the unloading phase, likely due to a thermal (rather than post-occlusive) hyperaemic response [note: the estimated post-occlusive thermal hyperaemia component in the SCI cohort is greater (i.e. ∼219%) than that of peak hyperaemia (i.e. ∼65%)]. Whilst our conservative approach to mechanical loading in the SCI cohort may have resulted in a small(-er) mechanical insult, the temperature-mediated changes in SkBF observed indicated that this group is likely to respond to more pronounced levels of mechanical loading, in combination with cooling, with a pattern similar to that observed in our healthy groups. Previous work in SCI participants has indeed highlighted that lowering skin temperature (i.e. ∼24 °C) during the unloading phase correlated with smaller reactive hyperaemia responses (Jan, 2020).

Despite reductions in post-occlusive reactive hyperaemia with cooling, we did not observe large changes in inflammatory biomarkers, nor did the combination of loading and heating (38 °C) elicit increases in inflammatory cytokines. We believe that this lack of responses was likely the result of the relatively mild mechanical insults used in this study. Indeed, despite the observed differences in perfusion, loading combined with local heating did not elicit increases in inflammatory cytokines, indicating that our experimental protocol was well tolerated and likely induced reversible vascular stress in all groups. Data from previous studies in rats (Lee *et al*., 2014) and mice (Wang *et al*., 2017) indicated that cooling decreases the inflammatory response to mechanical loading; yet those studies used much higher levels of loading (i.e. ∼700 mmHg) combined with longer exposures (i.e. up to 180 mins). Furthermore, it has been suggested that cold modulates the TNF-α expression via TRPM8 cold-sensitive neurones. In the context of pressure injuries, sustained elevation of TNF-α is typically present in chronic wound environments (Jiang *et al*., 2014), whereas acute and reversible loading may reflect reversible vascular stress without sufficient tissue injury and activation of TNF-α production. Similarly, IL-1β levels remained unchanged after loading, suggesting that healthy skin can withstand the experimental conditions and does not demand a large increase in leucocyte activity. Previous work has demonstrated that pressure-induced cytokine release can occur within 1 hour in in vitro models, although large quantities of IL-1α and IL-1RA remained within the tissue rather than diffusing into the sebum (Cornelissen *et al*., 2009; Jayabal *et al*., 2023a). Combined with the slow kinetics of sebaceous secretion, this may explain the negligible changes in cytokines observed in this study. Future studies should therefore consider expanding the boundaries of the mechanical insults applied to the skin (i.e. consider how loading can be applied in combination with shearing), and the associated time-dependent biomarker expressions (i.e. duration of insult and timing of sampling), beyond those investigated here, to better determine the potential immunological benefits of localised skin cooling.

### Clinical applications and translation

When taken together, empirical evidence from our 3 distinct participant cohorts indicated that locally cooling the skin to 24 and 16 °C, during and following mechanical loading, can largely suppress post-occlusive microvascular responses associated with increased PU-risk, in both healthy and vulnerable skin. Furthermore, the application of both cooling levels remained acceptable by most participants (>70%) throughout the trials. By combining physiological and perceptual readouts, the evidence from study could therefore inform the selection of design parameters for interventions and medical devices (e.g. support surfaces, dressings, adhesives etc.) that implement cooling technology to preserve skin tissue viability.

For example, our data indicate that when actively cooling the skin, lowering temperature to 24 °C may be preferable to 16 °C, as this approach would provide the same physiological and perceptual responses, but with a likely lower energy/power demand. This may allow for more energy-efficient approaches to local cooling technology (Arens *et al*., 2023). Furthermore, our data indicated that the benefits of cooling post-loading were likely contingent upon the continuous cooling of the unloaded skin tissue. This may therefore require consideration of wearable interventions/devices, by which the local cooling is maintained directly onto the skin, regardless of whether the individual unloads and/or repositions the tissue (e.g. consider bedridden patients who may be repositioned to unload skin areas, which may in turn lose contact with a cooling surface). Future translations of these findings into cooling technologies will also require consideration of how chronic cooling exposures may modulate the mechanical tolerance of skin tissue under daily living conditions.

#### Conclusions

We conclude that locally cooling the skin to 24 °C and 16 °C, during and following mechanical loading of the sacral area, can largely suppress post-occlusive microvascular responses associated with increased PU-risk, in both healthy younger and older adults, and in spinal cord injury patients. Furthermore, the application of both cooling levels remained acceptable by most participants (>70%) throughout the trials. As such, we concluded that lowering skin temperature to 24 °C during both loading and unloading may be preferable to 16 °C, as this approach would provide the same physiological and perceptual responses. By combining physiological and perceptual readouts, the evidence from this study could therefore inform the selection of design parameters for interventions and medical devices (e.g. support surfaces, dressings, adhesives etc.) that implement cooling technology to preserve skin tissue viability.

## Data Availability

All data produced in the present study are available upon reasonable request to the authors

